# Prevalence of malaria and associated factors among febrile under five children in Bora district, East Shoa zone, Oromia, Ethiopia, 2024: Institutional based cross sectional study

**DOI:** 10.1101/2025.05.04.25326951

**Authors:** Tsegaye Nigusu Alemu, Aman Yesuf Endries, Takele Gezahegn Demie, Dulelework Bayessa Hailemariam

## Abstract

**Introduction:** Malaria remains a major public health importance disease, especially, among under five children in Ethiopia, who were vulnerable group due to not well developed immunity. Although studies in regard to the burden of malaria among under-five children were available elsewhere, study in the low malaria transmission areas were scarcely available.

**Objective:** To determine the prevalence of malaria and identify associated factors among febrile under-five children in Bora district, East Shoa zone, Oromia region, Ethiopia in 2024.

**Methods:** An institution based cross sectional study was conducted among 466 children aged 6 to 59 months with febrile who visited health centers in Bora district from July 08 to August 09, 2024. The study participants were selected from each health centre considering proportionally allocation of the sample based on the number of febrile cases screened for malaria during the same period of the previous year. Data were collected using structured questionnaire and kobo collect version 2024.1.3, and cleaned then exported to STATA version 17 for analysis. Descriptive statistics was computed and reported by frequency and percentages. Bi-variable and multi-variable binary logistic regression analyses were employed to identify the factors associated with malaria. Thereafter, variable with Adjusted Odds Ratio(AOR) at 95% CI with P-value <0.05 was considered statistically significant variable.

**Results:** A total of 466 under five children were included in this study giving a response rate of 100% and the overall prevalence of malaria was 12.23%. This study showed under five children who lived in a home close to irrigation area(AOR=2.53, 95% CI: 1.15, 5.58), children who stayed outside during night(AOR=2.56, 95% CI: 1.21, 5.43) and children who did not wear a protective cloth at night(AOR=4.14, 95% CI: 1.03, 16.65) were significantly associated with the development of malaria.

**Conclusions:** Malaria remains with high prevalence among under five children. Children who lived in a home close to irrigation area, stayed outside during night and did not wear protective cloth during night were higher risk of developing malaria. Therefore, management of temporary stored water at irrigation area, not to stay outside during night and wearing protective cloth at night would reduce malaria among under-five children.

## Introduction

Malaria is an acute febrile illness caused by a parasite known as Plasmodium(1). The disease is transmitted via the bites of infected mosquitoes(2). It was one of the most severe public health problems worldwide and occurs mostly in poor tropical and subtropical areas of the world. Globally malaria was risk in 85 malaria endemic countries(3). In many of the countries affected by malaria, it was a leading cause of illness and death(4). In areas with high transmission, the most vulnerable groups were young children, who have not developed immunity to malaria yet, and pregnant women, whose immunity has been decreased by pregnancy(5).

According to World Health Organization (WHO) report, there were 249 million cases of malaria in 2022 compared to 244 million cases in 2021, an increase of five million cases and Ethiopia contributes for 1.3 million(32%) for these cases increment. The estimated number of malaria deaths stood at 608,000 in 2022 compared to 610,000 in 2021. African continues to carry a disproportionately high share of the global malaria burden. In 2022 the continent accounted for about 94% of all malaria cases and 95% of deaths. Children under five years of age accounted for about 78% of all malaria deaths in the continent(3).

There are five Plasmodium parasite species that cause malaria in humans and two of these species Plasmodium falciparum(P.F) and Plasmodium vivax(P.V) – pose the greatest threat. Symptoms of malaria can be mild or severe, usually appearing between 10 and 15 days after the mosquito bite. Mild symptoms include fever, headache, backache, joint pains and vomiting. Severe symptoms include extreme tiredness and fatigue, impaired consciousness, multiple convulsions, difficulty breathing, dark or bloody urine, jaundice(yellowing of the eyes) and abnormal bleeding(6).

In Ethiopia about three quarters(75%) of the country’s landmass was malaria endemic and about 52% of the total population was at risk of malaria infection(7). Malaria remains to be one of the major public health challenges in the country which shows marked seasonal, inter-annual and spatial variability due to large differences in climate(temperature, rainfall and relative humidity), topography(altitude, surface hydrology, vegetation cover and land use) and human settlement and population movement patterns that influence malaria risk(2).

P. falciparum and P. vivax were the two dominant parasite species causing malaria in Ethiopia, with relative frequencies of about 60% and 40%, respectively. P. malariae and P. ovale were rare and account for <1% of all confirmed malaria cases. The major malaria vector incriminated in Ethiopia was Anopheles Arabiensis; in some areas Anopheles Pharoensis, Anopheles Funestus and Anopheles Nili also play minor role in transmission of malaria. Anopheles Stephensi was a recently introduced malaria vector in Ethiopia(8).

Malaria can be treatable and preventable. A key interventions to control malaria include: prompt diagnosis and effective treatment with appropriate anti-malarial drugs, use of insecticidal bed nets; and indoor residual spraying of houses with insecticides to control the vector population(3). Early diagnosis and treatment are essential for more favorable malaria outcomes. As fever is a key manifestation of malaria in children, care-seeking for febrile children is crucial to reducing child morbidity and mortality. Whilst fever is an indicator of malaria in children, it can also be a sign of other acute infections(5).

The World Health Organization’s Global Technical Strategy for Malaria 2016-2030 has been developed with the aim of helping countries reduce the human suffering caused by the world’s deadliest mosquito-borne disease(9). Likewise, Ethiopia national malaria elimination strategic plan proposes to eliminate malaria in districts with an annual parasite index less than 10 by 2025 and the total elimination of malaria from Ethiopia by 2030(10). Bora district in which all populations in the district at risk of malaria was among the district targeted for malaria elimination.

Previous studies, focused only on malaria among adult, and malaria among under five children in area where malaria transmission were high. In areas with low malaria transmission, there was little evidence on the prevalence of malaria and related variables in children under five who were feverish. Despite the fact that health facilities reported malaria as one of the most common diseases in the community, there was limit evidence on the prevalence of malaria and related factors among febrile children under five who visited health facilities. Additionally, although the country had set targets that led to eliminate malaria through the involvement of vulnerable populations, the illness continues to be a public health concern in the area. Therefore, the aim of this study was to fill this gap, and to determine the prevalence of malaria and identify associated factors among febrile under-five children in Bora district.

## Materials and methods

### Study area, period and design

An institutional based cross sectional study design was employed in Bora district from July 07 to August 09, 2024. Bora district is one of the eleven districts found in East Shoa zone, Oromia regional state, Ethiopia. It is located in the Great Rift Valley at about 120Km distance East of Addis Ababa and 58Km from Adama, zonal town on the main road of Modjo to Hawasa. It is bordered in North by Lume district, in South by Dugda district, in East by Aris zone, and in West by Liban Chukala district.

Administratively the district is divided into 20 kebeles(18 rural and two urban) (kebele is Ethiopia’s lowest administrative unit), with the district administrative centre being at Bote (Alemtena) town. Majority of kebeles (90%) located in the rural area and all kebeles are malaria endemic. According to the 2007 national census, the projected population of the district was estimated to be 93,994 by 2024. Majority(80.3%) of the population in the district was rural dwellers and half (50.09%) of them was females in 2024(11).

The district had no hospital but it had three health centers and eighteen health posts providing several health services including prevention and control of malaria. The physical primary health service coverage of the district was above 96% and the district had 164 health professionals including health extension workers. Regarding malaria among under five children, the district health office report of 2023 shown that, there were 378 confirmed cases by microscope/RDT, zero clinically treated malaria cases and zero malaria death(11).

### Population

The source populations included all children age 6 to 59 months who have visited the health centers in Bora district. The study populations included all children age 6 to 59 months with fever who visited health centers in district during the study period.

### Eligibility criteria

Children aged 6 to 59 months with fever, live in the study area at least for six months and whose parents or caregivers signed a written consent form were included in the study. Children aged 6 to 59 months who were receiving anti-malarial treatment during data collection period were excluded.

### Sample size determination

First, we calculated the sample size for the first specific objective, considering the prevalence of malaria among under-five children using Epi Info version 7.2 for population survey considering the assumptions of 95% confidence level and 5% margin of error. Secondly, we calculated the sample size for the second specific objective considering factors associated with malaria among under-five children. Thus, we used double population proportion formula to calculate the sample size using Epi Info version 7.2 for cross sectional study considering the assumptions of 95% CI and 80% power. Finally, we considered the largest sample size, which was **424** among the calculated sample sizes for the second specific objective considering factors associated with malaria among under-five children from the previous study conducted in South Gondar zone(12). Considering 10% non-response rate, the final sample size was **466**.

### Sampling technique and procedures

All three health centers found in the district were purposively included in the study. Then, the number of study participants to be enrolled at each health centre were allocated for each health centre proportionally based on the number of febrile under-five children screened for malaria at each health centre during the same period of the previous year(July 07 to August 09, 2023) which was 735 cases. Thus, mathematically, number of febrile under-five children screened for malaria in each health centre during the same period of the previous year, multiplied by the total required sample size (N=466) then divided by the total sum of number of febrile cases visit in all the health centers(735 cases) as indicated in Figure 1 below.

Finally, systematic random sampling technique was applied to select study participants at each health centre. Thus, under-five children with febrile who visited the health centre laboratory for blood film examination and met inclusion criteria was recruited every k^th^ interval, considering the flow of cases in the health centre the previous year. The k^th^ interval was calculated by divide a total febrile under-five children screened for malaria during the same period of the previous year(735 cases) by the total required sample size(N=466) gave 1.57 ≈ 2. Therefore, the study participants who met the inclusion criteria were picked every two time after randomly selecting the first child.

**Figure 1:**
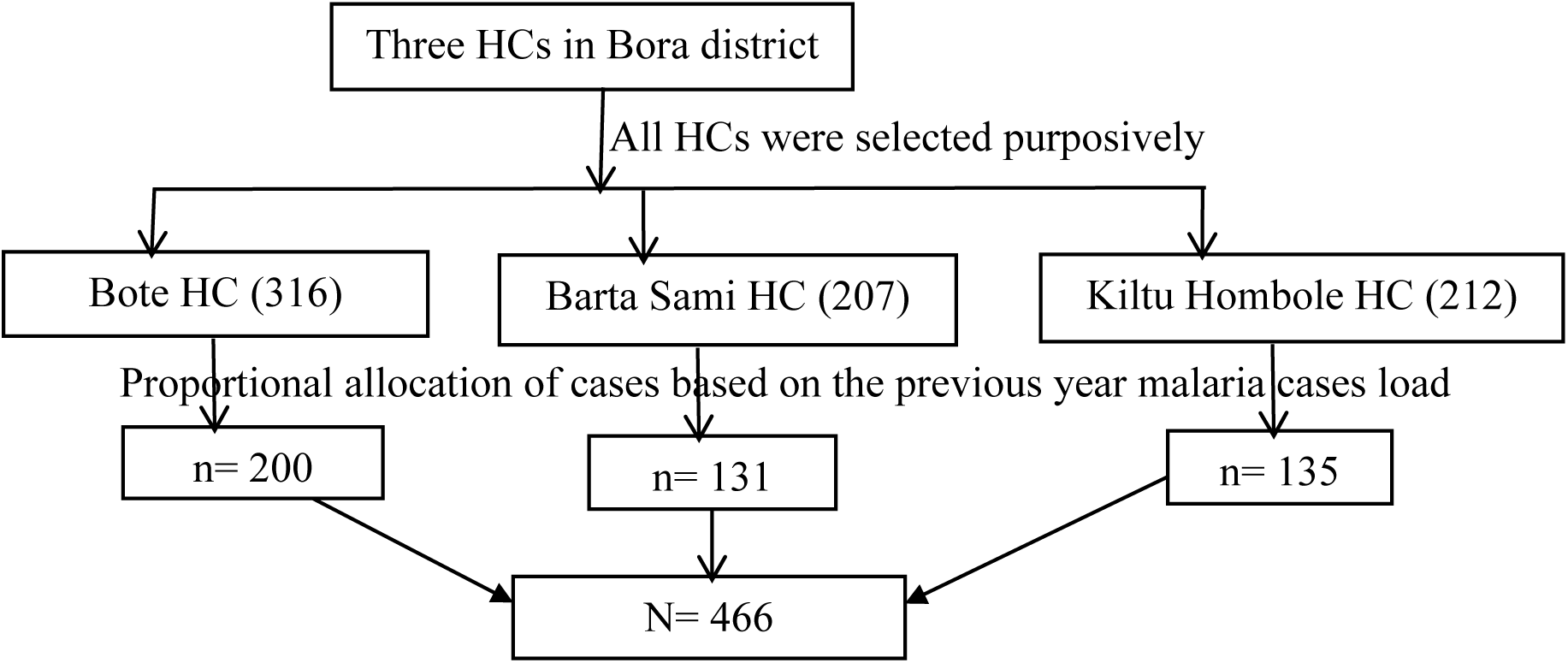
Schematic representation of sampling procedures for the study of prevalence of malaria and associated factors among under-five children in Bora district, July 2024

### Study variables

#### Dependent variable: Malaria status

Independent variables include: Socio-demographic factors such as age, sex, residence, religion, marital status, family size, number under five children, occupational status and educational status; Household and Environmental factors such as house door have screening, presence of a window, house windows have screening, wall of house have a hole, presence of mosquito breeding site such as old tires/broken glass/plastic container, plant in the containers/flower-pots, water pools, open shallow well, gutters to collect rainwater, stagnant water, intermittent river and irrigation, and Behavioral and knowledge related factors such as stay outside during night, wear a protective cloth at night, use of mosquito repellents, presence of sick person with symptom of fever in the house, history of travel to malaria endemic area, presence of sick person with symptom of fever in the house where you travel, presence of LLIN in the house, number of LLIN in the house, frequency of LLIN utilization, presence of a hole on LLIN, frequency of washing LLIN, house sprayed insecticidal residual spraying(IRS), time of IRS sprayed, knowledge of malaria transmission from person to person, knowledge of vectors that transmit malaria, knowledge of time of mosquito bite, knowledge of sign and symptoms of malaria, knowledge of treatability of malaria, and knowledge of malaria prevention and control methods.

### Operational definitions

- **Febrile**: refers high axillary temperature (≥37.5°C) or history of fever during 24 hrs proceeding the day of blood sampling
- **Door/window screening**: refers to any clothes or glasses on the door/window that protect mosquito not to enter the house
- **House wall hole**: this refers to any visible crack or crevice on the wall of the house through which mosquitoes could enter
- **Knowledge of sign and symptoms of malaria**: refers to fever, headache and back pain, chilling and shivering, nausea and loss of appetite, muscle and joint pain, vomiting, and thirsty. If the respondent stated that four and above “yes” among the above sign and symptoms of malaria he/she was considered as “Good knowledge of sign and symptoms of malaria and other ways he/she was considered as “poor knowledge
- **Knowledge of malaria prevention and control methods**: refers to utilization of bed nets, wearing long sleeved clothes, use repellents, close windows and doors at night, spraying insecticide chemicals, trimming bushes around the house and cleaning stagnant water around the house. If the respondent stated that four and above “yes” among the above malaria prevention and control methods he/she was considered as “Good knowledge of malaria prevention and control methods and other ways he/she was considered as “poor knowledge of malaria prevention and control methods
- **Prevalence of malaria**: refers to the percentage of the number of malaria positive individuals divided by the total malaria examined individuals
- **Presence of LLIN**: households having one or more LLIN during data collection period
- **Presence of a hole on LLIN**: refers to any tear on LLIN through which mosquitoes could enter
- **Presence of mosquito breeding site close to the home:** refers to presence of potential mosquito breeding site such as old tires/broken glass/plastic container, plant in the containers/flower-pots, water pools, open shallow well, gutters to collect rainwater, stagnant water, intermittent river and irrigation within 0.5Km radius from the home
- **Utilization of LLIN**: refers an individual’s reported regularly sleeping under LLIN within the past two-weeks before the onset of fever
- **Stagnant water**: It is a water body that may include, rainwater, ponds, natural water bodies, and standing water that remains after rainfall
- **Travel history**: refers to history of travel to known malaria endemic area two weeks before data collection
- **Wear protective cloth at night**: refers to children who wore long cloth to protect their leg and hand during night

### Data collection tool, methods and procedures

The primary source of data was collected at health centre on face-to-face interview administered using structured questionnaires prepared in English and translated to local language(Afan Oromo). Kobo collect version 2024.1.3 mobile application was used for the data collection. First, among children visited under five OPD, all children with fever were identified and referred to laboratory for slide test. Then medical laboratory technician of the respective health center collected capillary blood specimen from finger prick or big toe aseptically using sterile blood lancet to prepare thick and thin blood film smears to determine presence or absence of malaria parasites and identify the types of plasmodium species if malaria parasite present. Finally, data were collected from parents/caregivers by three level IV nurses who were selected based on their interest and experience on data collection. One supervisor(Health officer) and principal investigator were supervised the overall data collection process. The questionnaire gathers three groups of participants’ characteristics: socio-demographic related factors, household and environmental related factors, and behavioral and knowledge related factors.

### Blood specimen collection and processing

Capillary blood specimen was collected from finger prick or big toe aseptically using sterile blood lancet to prepare thick and thin blood film smears. Medical laboratory technician of the respective health centre was doing blood specimen collection while children attending the health centre due to febrile illness. In brief, the smear was air-dried. Thin film was fixed with methanol, and both thin and thick films was stained with 10% Giemsa stain for 15 minutes. All dried slides placed in slides boxes and was examined by laboratory technicians at the health centre laboratory.

The presence of malaria parasites on thick blood smear and the identification of Plasmodium species from smear was done, through oil immersed objective (100×), at 1000×magnification. The thick smear was used to determine whether the malaria parasite was present or absent and thin smear was used to identify the type of Plasmodium species. During the microscopic examination, a slide was regarded as negative after 200 fields were examined without finding of Plasmodium parasite by two laboratory technicians.

### Data quality control

The questionnaire was modified from previous literatures to the current context of the study based on the conceptual framework and study variables; and first prepared in English language and translated to ‘Afan Oromo’ which is the local language at the study area and re-translated back to English language for consistency. Data collectors (three level IV nurses) and supervisor (one Health Officer) were selected based on their interest and experience on data collection and supervision respectively. Then, they were trained for two days on the purposes and objectives of the study, logical order of questionnaire, data collection procedures, their roles in the data collection, how to ensure data quality, and importance of confidentiality and supervision.

Additionally, they were performing practical exercises to be familiar with the questionnaire by the investigators. Before the actual data collection, the questionnaire was pre-tested on 5% of the total sample size at Lume district (adjacent district) a week before commencement of the actual data collection. Based on the pre-test, a questionnaire was corrected to ensure clarity, wording, and logic sequence and skip patterns.

Supervision, follow-up and appropriate corrections were done on daily basis during the data collection process by the supervisor who was under the whole supervision of the principal investigator. Missing value, consistency and completeness of data were checked and cleaned. If any error was encountered it was addressed immediately. To ensure the quality of the microscopic examinations, all positive and 10% of the negative slides were re-examined by the third reader (internal quality assurance) to remove discrepant result.

### Data processing and analysis

Prior to entering the data into the computer, missing value, consistency and its completeness was checked and data were cleaned. Then, the data were exported from kobo collect version 2024.1.3 to STATA version 17 for further analysis. The outcome variable (malaria status) was categorized as “Positive or negative” and displayed by frequency and percentage using tables/graphs/charts, and text descriptions accordingly

The normality of continuous variable was checked using Shapiro-Wilks test with p-value >0.05 considered as normal distribution. Descriptive statistics were computed for normal distribution continuous variables and reported as mean with standard deviation. For non-normal distribution continuous variables median with inter-quartile range(IQR) were reported. Categorical variable was reported by frequency and percentages.

Whether the necessary assumption was fulfilled for the application of binary logistic regression was checked(outcome variable with two responses). Both bi-variable and multi-variable binary logistic regression analysis were performed to see the association between dependent and independent variables using binary logistic regression. Variables in bi-variable binary logistic regression analysis with p-value < 0.25 were entered into multi-variable binary logistic regression analysis after multi-collinearity and model goodness of fit test were checked.

Multi-collinearity test was checked using variance inflation factor (VIF) to identify collinearity between the independent variables. The model goodness of fit test waschecked by Hosmer-Lemeshow goodness of fit test for the final model and; chi-square and the p-value of the model fitness of the test were identified. In this regard, the Hosmer-Lemeshow’s goodness of fit test with large p-value (p>0.05) was checked to see good fitness. After multi-collinearity and model goodness of fit was checked, variables without multi-collinearity(VIF less than five) and with good fit model was entered into multi-variable binary logistic regression analysis.

Finally, multi-variable binary logistic regression analysis was used for controlling confounding factors and to identify significant determinants of malaria infection among under-five children. At the end, variable with Adjusted Odds Ratio(AOR) at 95% CI, P-value <0.05 was considered statistically significance variable and presented by tables/graphs/charts, and text descriptions accordingly.

### Ethical considerations

The proposal was reviewed and approved by the Institutional Review Board (IRB) of St. Paul’s Hospital Millennium Medical College(SPHMMC). Support letter was obtained from SPHMMC School of public health and Bora district health office. The purpose of the study was explained to each of the study participants. Participants were informed that their participation was on a voluntary basis. Data collection was begun after written informed assent was obtained from each parent/caregiver of the children. To make sure privacy and confidentiality of participants information; anonymous typing was used instead of their names and no participants identifiers was written on the questionnaire. To keep the privacy of the participants, all participants were interviewed alone. The confidentiality of information obtained from the interviewee was securely kept throughout the entire study period. During each data collections process, for those children who were found malaria positive, free treatment was provided at health facility.

## Results

### Socio-demographic characteristics

In this study, a total of 466 participants were participated giving a response rate of 100% recorded from their parents or caretakers. The age of the participants ranged 6-57 months and age was not normally distributed, thus the median age was 24.00(IQR: 24.00) months. More than a quarter(26.6%) were age 12-23 months. More than half(51.5%) of the study participants were male. More than two third(66.95%) of the study participants were rural residents.

More than a quarter(29.6%) of the respondents were protestant in religion while more than three fourth(80.5%) married in marital status. On the other hand, more than half(54.3%) and nearly two third(63.5%) of the respondents had less than five family size and had only one under five children in the house respectively. With regard to occupation and educational status of the respondents; about half(50.6%) were housewife and nearly one third(32.6%) could read and write(Table 1).

**Table 1:**
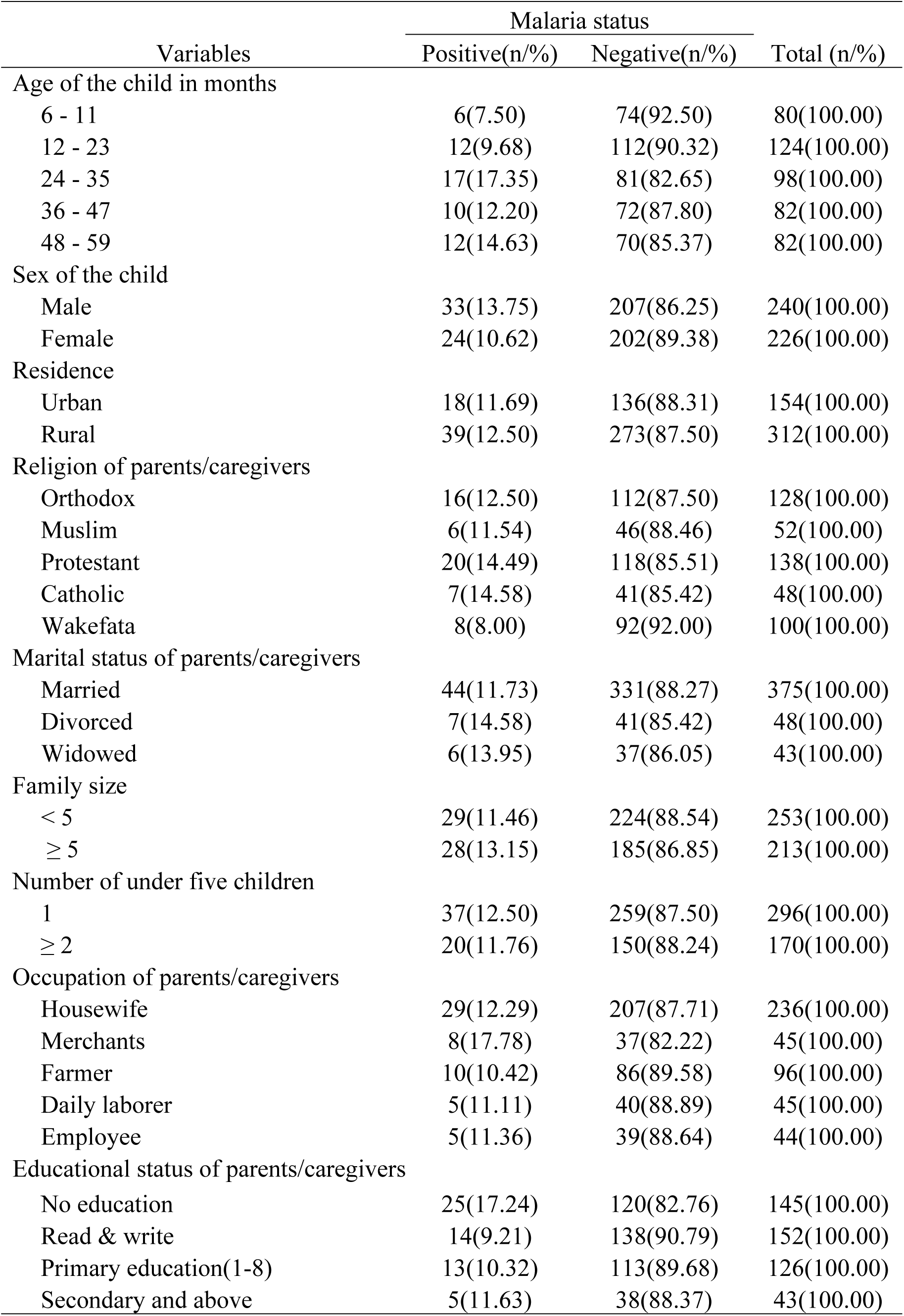
Socio-demographic characteristics of study participants in Bora district, East Shoa zone, Oromia, Ethiopia, July 2024 (n = 466)

| Variables | Malaria status |  | Total (n/%) |
| --- | --- | --- | --- |
|  | Positive(n/%) | Negative(n/%) |  |
| Age of the child in months |  |  |  |
| 6 - 11 | 6(7.50) | 74(92.50) | 80(100.00) |
| 12 - 23 | 12(9.68) | 112(90.32) | 124(100.00) |
| 24 - 35 | 17(17.35) | 81(82.65) | 98(100.00) |
| 36 - 47 | 10(12.20) | 72(87.80) | 82(100.00) |
| 48 - 59 | 12(14.63) | 70(85.37) | 82(100.00) |
| Sex of the child |  |  |  |
| Male | 33(13.75) | 207(86.25) | 240(100.00) |
| Female | 24(10.62) | 202(89.38) | 226(100.00) |
| Residence |  |  |  |
| Urban | 18(11.69) | 136(88.31) | 154(100.00) |
| Rural | 39(12.50) | 273(87.50) | 312(100.00) |
| Religion of parents/caregivers |  |  |  |
| Orthodox | 16(12.50) | 112(87.50) | 128(100.00) |
| Muslim | 6(11.54) | 46(88.46) | 52(100.00) |
| Protestant | 20(14.49) | 118(85.51) | 138(100.00) |
| Catholic | 7(14.58) | 41(85.42) | 48(100.00) |
| Wakefata | 8(8.00) | 92(92.00) | 100(100.00) |
| Marital status of parents/caregivers |  |  |  |
| Married | 44(11.73) | 331(88.27) | 375(100.00) |
| Divorced | 7(14.58) | 41(85.42) | 48(100.00) |
| Widowed | 6(13.95) | 37(86.05) | 43(100.00) |
| Family size |  |  |  |
| < 5 | 29(11.46) | 224(88.54) | 253(100.00) |
| ≥ 5 | 28(13.15) | 185(86.85) | 213(100.00) |
| Number of under five children |  |  |  |
| 1 | 37(12.50) | 259(87.50) | 296(100.00) |
| ≥ 2 | 20(11.76) | 150(88.24) | 170(100.00) |
| Occupation of parents/caregivers |  |  |  |
| Housewife | 29(12.29) | 207(87.71) | 236(100.00) |
| Merchants | 8(17.78) | 37(82.22) | 45(100.00) |
| Farmer | 10(10.42) | 86(89.58) | 96(100.00) |
| Daily laborer | 5(11.11) | 40(88.89) | 45(100.00) |
| Employee | 5(11.36) | 39(88.64) | 44(100.00) |
| Educational status of parents/caregivers |  |  |  |
| No education | 25(17.24) | 120(82.76) | 145(100.00) |
| Read & write | 14(9.21) | 138(90.79) | 152(100.00) |
| Primary education(1-8) | 13(10.32) | 113(89.68) | 126(100.00) |
| Secondary and above | 5(11.63) | 38(88.37) | 43(100.00) |

### Prevalence of malaria among under-five children

Among 466 study participants with fever visited health facilities and tested for malaria parasite by microscopy, 57 were positive for malaria and result in 12.23% prevalence of malaria among under-five children. Among those who tested positive for malaria, more than three fourth(77.2%) were infected with P.F species (Figure 2).

**Figure 2:**
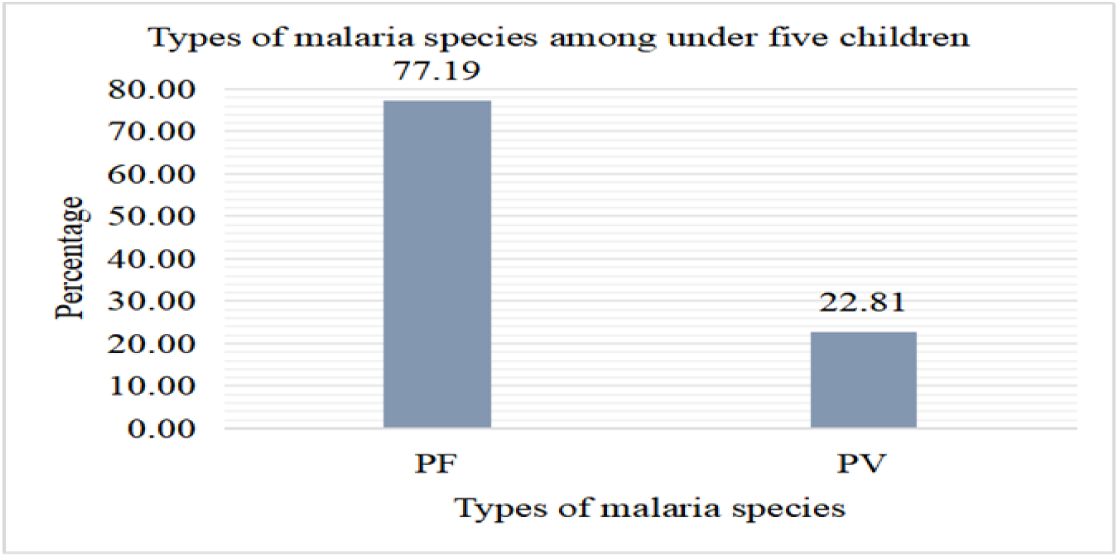
Distribution of malaria species among febrile under five children in Bora district, East Shoa zone, Oromia, Ethiopia, July 2024 (n = 57)

On the other hand, among those who tested positive for malaria, more than a quarter(29.8%) were aged 24-35 months. With respect to sex and resident of malaria positive, more than half(57.9%) were male in sex and more than two third(68.4%) were rural residents(Table 2).

**Table 2:** Malaria species distribution by age, sex and residence among febrile under five children in Bora district, East Shewa zone, Oromia, Ethiopia, July 2024 (n = 57)

| Variables | Total (n/%) | Malaria<br>positive(n/%) | Malaria species |  |
| --- | --- | --- | --- | --- |
|  |  |  | PF(n/%) | PV(n/%) |
| Age of the child in months |  |  |  |  |
| 6 - 11 | 80(17.17) | 6(10.53) | 5(11.36) | 1(7.69) |
| 12 - 23 | 124(26.61) | 12(21.05) | 8(18.18) | 4(30.77) |
| 24 - 35 | 98(21.03) | 17(29.82) | 16(36.36) | 1(7.69) |
| 36 - 47 | 82(17.60) | 10(17.54) | 7(15.91) | 3(23.08) |
| 48 - 59 | 82(17.60) | 12(21.05) | 8(18.18) | 4(30.77) |
| Sex of the child |  |  |  |  |
| Male | 240(51.50) | 33(57.89) | 29(65.91) | 4(30.77) |
| Female | 226(48.50) | 24(42.11) | 15(34.09) | 9(69.23) |
| Residence of the child |  |  |  |  |
| Urban | 154(33.05) | 18(31.58) | 13(29.55) | 5(38.46) |
| Rural | 312(66.95) | 39(68.42) | 31(70.45) | 8(61.54) |

### Household and environmental related characteristics

Nearly one third(30.3%) of the study participants had house with screened door and more than three fourth(77.5%) had house with screened window. Majority(89.5%) of the study participants wall of the house did not have visible hole. Regarding the presence of potential mosquito breeding site, only 43(9.2%) and 54(11.6%) of the study participants were living a home/house with water in broken container and close to irrigation respectively(Table 3).

**Table 3:** Household and environmental related characteristics by malaria status among febrile under five children in Bora district, East Shoa zone, Ethiopia, July 2024 (n = 466)

| Variables | Malaria status |  | Total (n/%) |
| --- | --- | --- | --- |
|  | Positive(n/%) | Negative(n/%) |  |
| House door have screened |  |  |  |
| Yes | 16(11.35) | 125(88.65) | 141(100.00) |
| No | 41(12.62) | 284(87.38) | 325(100.00) |
| House have window |  |  |  |
| Yes | 23(10.55) | 195(89.45) | 218(100.00) |
| No | 34(13.71) | 214(86.29) | 248(100.00) |
| House window have screened |  |  |  |
| Yes | 16(9.47) | 153(90.53) | 169(100.00) |
| No | 7(14.29) | 42(85.71) | 49(100.00) |
| Wall of house have visible hole |  |  |  |
| Yes | 7(14.29) | 42(85.71) | 49(100.00) |
| No | 50(11.99) | 367(88.01) | 417(100.00) |
| Presence of mosquito breeding site |  |  |  |
| Water in broken container |  |  |  |
| Yes | 9(20.93) | 34(79.07) | 43(100.00) |
| No | 48(11.35) | 375(88.65) | 423(100.00) |
| Plant in the containers /flower-pots |  |  |  |
| Yes | 5(8.77) | 52(91.23) | 57(100.00) |
| No | 52(12.71) | 357(87.29) | 409(100.00) |
| Water pools |  |  |  |
| Yes | 30(16.48) | 152(83.52) | 182(100.00) |
| No | 27(9.51) | 257(90.49) | 284(100.00) |
| Open shallow well |  |  |  |
| Yes | 15(13.27) | 98(86.73) | 113(100.00) |
| No | 42(11.90) | 311(88.10) | 353(100.00) |
| Gutters to collect rainwater |  |  |  |
| Yes | 12(9.02) | 121(90.98) | 133(100.00) |
| No | 45(13.51) | 288(86.49) | 333(100.00) |
| Stagnant water |  |  |  |
| Yes | 40(12.50) | 280(87.50) | 320(100.00) |
| No | 17(11.64) | 129(88.36) | 146(100.00) |
| Intermittent river |  |  |  |
| Yes | 6(13.33) | 39(86.67) | 45(100.00) |
| No | 51(12.11) | 370(87.89) | 421(100.00) |
| Irrigation |  |  |  |
| Yes | 15(27.78) | 39(72.22) | 54(100.00) |
| No | 42(10.19) | 370(89.81) | 412(100.00) |

### Behavioral and knowledge related characteristics

More than one third(34.3%) of the participants stayed outside during night while only 66(14.2%) of children wear protective cloths during night. All of the participants did not use mosquito repellent and only 44(9.4%) had history of travel to malaria endemic area in the past one month before data collection. Slightly less than two third(62.9%) of the participants had LLIN and 32.0% had only one LLIN. With respect to frequency of LLIN use, only 45(9.7%) of the study participants were using regularly. On the other hand, slightly less than two third(62.7%) of the respondents knew malaria was transmitted from person to person. More than three fourth(76.8%) of the respondents respond mosquito bite during night time(Table 4).

**Table 4:** Behavioral and knowledge related characteristics and malaria status among febrile under five children in Bora district, East Shewa zone, Oromia, Ethiopia, July 2024 (n = 466)

| Variables | Malaria status |  | Total (n/%) |
| --- | --- | --- | --- |
|  | Positive(n/%) | Negative(n/%) |  |
| Stay outside during night |  |  |  |
| Yes | 29(18.13) | 131(81.88) | 160(100.00) |
| No | 28(9.15) | 278(90.85) | 306(100.00) |
| Wear a protective cloth at night |  |  |  |
| Yes | 3(4.55) | 63(95.45) | 66(100.00) |
| No | 54(13.50) | 346(86.50) | 400(100.00) |
| Use mosquito repellents |  |  |  |
| No | 57(12.23) | 409(87.77) | 466(100.00) |
| Presence of sick person in the house |  |  |  |
| Yes | 3(6.82) | 41(93.18) | 44(100.00) |
| No | 54(12.80) | 368(87.20) | 422(100.00) |
| Travel history to malarious area |  |  |  |
| Yes | 6(13.64) | 38(86.36) | 44(100.00) |
| No | 51(12.09) | 371(87.91) | 422(100.00) |
| presence of sick person at the place where you have been |  |  |  |
| No | 6(13.64) | 38(86.36) | 44(100.00) |
| Presence of LLIN in the house |  |  |  |
| Yes | 33(11.26) | 260(88.74) | 293(100.00) |
| No | 24(13.87) | 149(86.13) | 173(100.00) |
| Number of LLIN |  |  |  |
| never has LLIN | 24(13.87) | 149(86.13) | 173(100.00) |
| 1 | 9(6.04) | 140(93.96) | 149(100.00) |
| 2 | 18(17.82) | 83(82.18) | 101(100.00) |
| ≥3 | 6(13.95) | 37(86.05) | 43(100.00) |
|  | Positive(n/%) | Negative(n/%) |  |
| Frequency of LLIN use |  |  |  |
| Regularly | 6(13.33) | 39(86.67) | 45(100.00) |
| Occasionally | 16(10.96) | 130(89.04) | 146(100.00) |
| Never use | 35(12.73) | 240(87.27) | 275(100.00) |
| Presence of a hole on LLIN |  |  |  |
| Yes | 7(15.22) | 39(84.78) | 46(100.00) |
| No | 26(10.53) | 221(89.47) | 247(100.00) |
| Frequency of washing LLIN |  |  |  |
| ≤ 2 | 6(13.04) | 40(86.96) | 46(100.00) |
| > 2 | 6(10.34) | 52(89.66) | 58(100.00) |
| Never wash | 21(11.11) | 168(88.89) | 189(100.00) |
| House sprayed with IRS in the last six months |  |  |  |
| No | 57(12.23) | 409(87.77) | 466(100.00) |
| Knowledge of malaria transmission |  |  |  |
| Yes | 31(10.62) | 261(89.38) | 292(100.00) |
| No | 26(14.94) | 148(85.06) | 174(100.00) |
| Knowledge of vectors that transmit malaria |  |  |  |
| Mosquito | 31(10.62) | 261(89.38) | 292(100.00) |
| Knowledge of duration of mosquito bite |  |  |  |
| During the day time | 5(10.00) | 45(90.00) | 50(100.00) |
| During night time | 45(12.57) | 313(87.43) | 358(100.00) |
| Any time | 7(12.07) | 51(87.93) | 58(100.00) |
| Knowledge common signs and symptoms of malaria |  |  |  |
| Fever |  |  |  |
| Yes | 48(15.29) | 266(84.71) | 314(100.00) |
| No | 9(5.92) | 143(94.08) | 152(100.00) |
| Headache and back pain |  |  |  |
| Yes | 47(14.46) | 278(85.54) | 325(100.00) |
| No | 10(7.09) | 131(92.91) | 141(100.00) |
| Chilling and shivering |  |  |  |
| Yes | 46(12.74) | 315(87.26) | 361(100.00) |
| No | 11(10.48) | 94(89.52) | 105(100.00) |
| Nausea and loss of appetet |  |  |  |
| Yes | 49(13.61) | 311(86.39) | 360(100.00) |
| No | 8(7.55) | 98(92.45) | 106(100.00) |
| Muscle and joint pain |  |  |  |
| Yes | 23(11.73) | 173(88.27) | 196(100.00) |
| No | 34(12.59) | 236(87.41) | 270(100.00) |

Table 4: (Continue)
| Variables | Malaria status |  | Total (n/%) |
| --- | --- | --- | --- |
|  | Positive(n/%) | Negative(n/%) |  |
| Muscle and joint pain |  |  |  |
| Yes | 23(11.73) | 173(88.27) | 196(100.00) |
| No | 34(12.59) | 236(87.41) | 270(100.00) |
| Vomiting |  |  |  |
| Yes | 44(13.10) | 292(86.90) | 336(100.00) |
| No | 13(10.00) | 117(90.00) | 130(100.00) |
| Thirsty |  |  |  |
| Yes | 54(15.93) | 285(84.07) | 339(100.00) |
| No | 3(2.36) | 124(97.64) | 127(100.00) |
| Knowledge of malaria prevent and control methods |  |  |  |
| Sleeping in bed nets |  |  |  |
| Yes | 55(14.44) | 326(85.56) | 381(100.00) |
| No | 2(2.35) | 83(97.65) | 85(100.00) |
| Wearing long sleeved clothes |  |  |  |
| Yes | 13(9.22) | 128(90.78) | 141(100.00) |
| No | 44(13.54) | 281(86.46) | 325(100.00) |
| Use mosquito repellent |  |  |  |
| Yes | 14(9.93) | 127(90.07) | 141(100.00) |
| No | 43(13.23) | 282(86.77) | 325(100.00) |
| Close windows and doors at night |  |  |  |
| Yes | 50(13.93) | 309(86.07) | 359(100.00) |
| No | 7(6.54) | 100(93.46) | 107(100.00) |
| Spraying insecticide chemicals |  |  |  |
| Yes | 55(14.82) | 316(85.18) | 371(100.00) |
| No | 2(2.11) | 93(97.89) | 95(100.00) |
| Trimming bushes around the house |  |  |  |
| Yes | 49(13.73) | 308(86.27) | 357(100.00) |
| No | 8(7.34) | 101(92.66) | 109(100.00) |
| Cleaning stagnant water around the house |  |  |  |
| Yes | 54(14.10) | 329(85.90) | 383(100.00) |
| No | 3(3.61) | 80(96.39) | 83(100.001) |

### Factors associated to malaria among under-five children

In bi-variable binary logistic regression analysis; variable such as age, religion, water in broken container, gutters to collect rainwater, stay outside during night, child did not wear a protective cloth at night, number of LLIN in the HH and knowledge of malaria transmission were associated with malaria among under five years children. Additionally, presence of water pools, presence of irrigation, knowledge common signs and symptoms of malaria, and knowledge of methods of malaria prevent and control were significantly associated with malaria among under five years children. However, in multi-variable binary logistic regression analysis; only variables such as presence of irrigation close to the home, stay outside during night and child did not wear a protective cloth at night were significantly associated with malaria among under five years children.

The odds of developing malaria was 2.53 times higher among under five years children who live in a house close to irrigation, compared to children who did not live in a house close to irrigation(AOR = 2.53, 95% CI: 1.15, 5.58). The odd of developing malaria was 2.56 times higher among under five years children who stayed outside during night, compared to those who did not stay outside during night(AOR = 2.56, 95% CI: 1.21, 5.43). With respect to wearing protective cloth at night, the odd of developing malaria was 4.14 times higher among under five years children who did not wear a protective cloth at night, compared to those who wear a protective cloth at night(AOR = 4.14, 95% CI: 1.03, 16.65) (Table 5).

The Hosmer-Lemeshow goodness of fit test was used to assess the fitness of the final model and the final model was fit well with chi-square(X^2^)= 9.85 and P value of 0.28.

**Table 5:** Bi-variable and multi-variable logistic regression analysis of factors of malaria among febrile under five children in Bora district, East Shoa zone, Oromia, Ethiopia, 2024.

| Variables | Positive | Negative | COR (95% CI) | P-value | AOR (95% CI) |
| --- | --- | --- | --- | --- | --- |
| **Age of the child in months** |  |  |  |  |  |
| 6 - 11 | 6 | 74 | 1.00 |  | 1.00 |
| 12 - 23 | 12 | 112 | 1.32(0.48, 3.68) | 0.59 | 1.00(0.33, 3.03) |
| 24 - 35 | 17 | 81 | 2.59(0.97, 6.92) | **0.06** | 2.13(0.74, 6.14) |
| 36 - 47 | 10 | 72 | 1.71(0.59, 4.96) | 0.32 | 1.01(0.28, 3.63) |
| 48 - 59 | 12 | 70 | 2.11(0.75, 5.94) | **0.16** | 1.66(0.49, 5.60) |
| **Religion** |  |  |  |  |  |
| Orthodox | 16 | 112 | 1.64(0.67, 4.01) | 0.28 | 1.11(0.40, 3.11) |
| Muslim | 6 | 46 | 1.50(0.49, 4.58) | 0.48 | 1.08(0.31, 3.69) |
| Protestant | 20 | 118 | 1.95(0.82, 4.62) | **0.13** | 1.50(0.56, 4.04) |
| Catholic | 7 | 41 | 1.96(0.67, 5.78) | **0.22** | 1.44(0.43, 4.87) |
| Wakefata | 8 | 92 | 1.00 |  | 1.00 |
| **Presence of mosquito breeding site** |  |  |  |  |  |
| **Water in broken container** |  |  |  |  |  |
| Yes | 9 | 34 | 2.07(0.94, 4.57) | **0.07** | 1.72(0.70, 4.28) |
| No | 48 | 375 | 1.00 |  | 1.00 |
| **Water pools** |  |  |  |  |  |
| Yes | 30 | 152 | 1.88(1.08, 3.28) | **0.03** | 1.67(0.85, 3.31) |
| No | 27 | 257 | 1.00 |  | 1.00 |
| **Gutters to collect rainwater** |  |  |  |  |  |
| Yes | 12 | 121 | 0.63(0.32, 1.24) | **0.18** | 0.77(0.34, 1.72) |
| No | 45 | 288 | 1.00 |  | 1.00 |
| **Irrigation** |  |  |  |  |  |
| Yes | 15 | 39 | 3.39(1.72, 6.66) | **0.00** | **2.53(1.15, 5.58)\*** |
| No | 42 | 370 | 1.00 |  | 1.00 |
| **Stay outside during night** |  |  |  |  |  |
| Yes | 29 | 131 | 2.20(1.26, 3.85) | **0.01** | **2.56(1.21, 5.43)\*\*** |
| No | 28 | 278 | 1.00 |  | 1.00 |
| **Wear a protective cloth at night** |  |  |  |  |  |
| Yes | 3 | 63 | 1.00 |  | 1.00 |
| No | 54 | 346 | 3.28(0.99, 10.81) | **0.05** | **4.14(1.03, 16.65)\*** |
| **Number of LLIN** |  |  |  |  |  |
| never has LLIN | 24 | 149 | 0.99(0.38, 2.61) | 0.99 | 2.24(0.70, 7.13) |
| 1 | 9 | 140 | 0.40(0.13, 1.18) | **0.13** | 1.94(0.51, 7.40) |
| 2 | 18 | 83 | 1.34(0.49, 3.64) | 0.57 | 1.74(0.57, 5.28) |
| >=3 | 6 | 37 | 1.00 |  | 1.00 |

Table 5: (Continue)
| Variables | Positive | Negative | COR (95% CI) | P-value | AOR (95% CI) |
| --- | --- | --- | --- | --- | --- |
| Knowledge of malaria transmission |  |  |  |  |  |
| Yes | 31 | 261 | 1.00 |  | 1.00 |
| No | 26 | 148 | 1.48(0.85, 2.59) | **0.17** | 0.93(0.48, 1.77) |
| Knowledge signs and symptoms |  |  |  |  |  |
| Good | 50 | 289 | 1.00 |  | 1.00 |
| Poor | 7 | 120 | 0.34(0.15, 0.76) | **0.01** | 0.40(0.16, 1.01) |
| Knowledge of prevent and control |  |  |  |  |  |
| Good | 54 | 309 | 1.00 |  | 1.00 |
| Poor | 3 | 100 | 0.17(0.05, 0.56) | **0.00** | 0.26(0.07, 1.00) |

**Significance level:** \* Significant ( $p < 0.05$ ), \*\*strongly significant ( $p < 0.01$ ), \*\*\*highly significant ( $p < 0.001$ ); **1.00:** reference variable

## Discussion

This study was conducted to determine the prevalence of malaria and identify associated factors among febrile under-five children in Bora district. The results of study showed that, a total of 466 participants were participated giving a response rate of 100%. The overall prevalence of malaria was 12.23%. About 11.6% of the study participants were living in a home close to irrigation area. More than one third(34.3%) stayed outside during night and majority(85.8%) did not wear protective cloth during night.

The study revealed that, higher prevalence of malaria among febrile under-five children who visited health facility. This was higher compared to the study conducted in Benishangul-Gumuz region, Ethiopia in 2021(3.9%)(13), in southern Ethiopia in 2023(7.8%)(14) and in Wogera district, Northwest Ethiopia in 2021(8.7%)(15) all showed that, lower prevalence of malaria among under five children.

On contrary, lower compared to the study conducted in sub-Saharan African countries in 2023(29.0%)(16), in Uganda in 2023(27.41%)(17) and in Nigeria in 2023(27.0%)(18) all showed that, higher prevalence of malaria among under five children. In similar way, lower compared to the study conducted in Tanzania in 2021(15.9%)(19), in Afar region, Ethiopia in 2019(64.0%) (20) and in Ziquala district, Northeast Ethiopia in 2022(24.6%)(21) all showed that, higher prevalence of malaria among under five children. The study also showed that lower compared to the study conducted in Sheko District, Southwest Ethiopia in 2023(23.4%)(22), in Arba-Minch Zuria, South Ethiopia in 2020(22.1%)(23), in Northeastern Ethiopia in 2023(15.5%)(24) and Kaffa zone, Southwest Ethiopia in 2022(13.6%)(25) that all showed, higher prevalence of malaria among under five children. This discrepancy might be as a result of difference in the study period, geographical area, and malaria prevention and control programs implementation in the study area.

The study has also showed that living in a house close to irrigation area was significantly associated with the higher odds of malaria among under-five children. This was supported by the study conducted in Southern Ethiopia(14), in Abergelle district, Tigray region, Ethiopia(26), in Bena Tsemay district, Southern Ethiopia(27) and in Simada district, Northwest Ethiopia(28) all showed that, the odds of developing malaria was higher among under five children who live in a house close to irrigation area. This might be due to presence of numerous collection of temporary stored water at irrigation site, which was a potential breeding site for mosquito and created suitable condition for reproduction of mosquito. This might also be due to mosquito can travel up to 7Km in wind direction and infecting people.

The study has also showed that staying outside during night in the past two weeks was significantly associated with the higher odds of malaria among under-five children. This study was supported by the study conducted in Wogera district, Northwest Ethiopia(15), in Benishangul-Gumuz region, Ethiopia(13) and in Ziway-Dugda district, Ethiopia(29) all showed that, the odds of developing malaria was higher among under five years children who stayed outside during night. The study also supported by the study conducted in Siraro district, Oromia region, Ethiopia(30) and Zimbabwe(31) both showed that, the odds of developing malaria was higher among under five years children who stayed outside during night. This might be due to the main biting time of female Anopheles mosquito that transmit malaria was at early night time and children who stay outside at this time had higher risk of exposure to outdoor feeding Anopheles mosquito. This might also be due to the study was conducted at the main agricultural season in the study area that people in rural area could stay late outside the home during night time without wearing protective clothes at night.

Additionally, the study also showed that, higher odds of developing malaria among under five years children who did not wear a protective cloth at night. This study was supported with the study conducted in Bena Tsemay district, Southern Ethiopia(27) and in Chipingo district, Zimbabwe(32) both showed that, the odds of developing malaria was higher among under five years children who who did not wear a protective cloth at night. On the other hand, the study conducted in Laely Adyabo district, Northern Ethiopia(33) and in Mudzi district, Zimbabwe(34) both showed that wearing long-sleeved clothes at night was protective from developing malaria. This might be due to children who did not wear mosquito bite protective cloth could be easily exposed to mosquito bite affected by malaria disease.

This study had its own strengths and limitations: The strength of this study was able to collect primary data directly from parents or caregivers rather than record review. The study also had limitations; the study based only on parents or caregivers response, however, some factors need physical observations (for example: house conditions, presence of LLIN, number of LLIN and utilization of LLIN). Additionally, in this study we did not look for factors such as HH water source, HH income and health seeking behavior of the study participants. Therefore, we suggest further study on these limitations.

## Conclusion

This study showed high prevalence of malaria among under-five children in the study area. Living in a home close to irrigation area, staying outside during night and not wearing protective cloth during night were identified associated factors of malaria among under-five.

## Data Availability

All relevant data are within the manuscript and its Supporting Information files.

## Ethics statement

The Ethical Committee that approved this study was the Institutional Review Board of St. Paul’s Hospital Millennium Medical College, School of Public Health. Data collection was begun after written informed assent was obtained from each parent/caregiver of the children. To make sure privacy and confidentiality of participants information; anonymous typing was used instead of their names and no participants identifiers was written on the questionnaire.

## Authors contributions

Conceptualization, TNA, AYE, TGD and DBH; data curation, TNA, AYE, TGD and DBH; formal analysis, TNA, AYE, TGD and DBH; funding acquisition, TNA; project administration, TNA; supervision, TNA; writing original draft, TNA, AYE, TGD and DBH; writing-review & editing, TNA, AYE, TGD and DBH. All authors contributed to the article and approved the submitted version.

## Acknowledgments

The authors like to acknowledge the study participants, data collectors, supervisors, colleagues, Bora district health office and St. Paul’s Hospital Millennium Medical College, School of Public Health.

